# Safety and feasibility of a combined neuromodulation and yoga intervention for individuals with mild traumatic brain injury and chronic pain: a pilot study

**DOI:** 10.1101/2023.12.01.23299142

**Authors:** Kelly Krese, Alexandra Aaronson, Kyla Z Donnelly, Rachana Shah, Sarmistha Chaudhuri, Sonia Bobra, Bella Etingen, Sabrina Bedo, Ibuola Kale, Andrea Billups, Kalea Colletta, Sandra Kletzel, Amy Kemp, Theresa L Bender Pape, Dulal Bhaumik, Amy A Herrold

**Affiliations:** Brain Innovation Center, Shirley Ryan AbilityLab, 355 E Erie St, Chicago, IL 60611, USA; Research and Development Service, Edward Hines Jr. VA Hospital, 5000 S 5 Ave, Hines, IL, 60141, USA; Chicago Association for Research and Education in Science, Edward Hines Jr. VA Hospital, 5000 S 5 Ave, Hines, IL, 60141, USA; Mental Health Service Line, Edward Hines, Jr. VA Hospital, 5000 S 5 Ave, Hines, IL, 60141, USA; Department of Psychiatry, University of Illinois at Chicago, 601 S Morgan St, Chicago, IL 60607, USA; LoveYourBrain Foundation, Norwich, VT USA; Department of Physical Medicine and Rehabilitation, Edward Hines, Jr. VA Hospital, 5000 S 5 Ave, Hines, IL, 60141, USA; Department of Radiology, Edward Hines, Jr. VA Hospital, 5000 S 5 Ave, Hines, IL, 60141, USA; Loyola University Chicago Stritch School of Medicine, 2160 S 1st Ave, Maywood, IL 60153, USA; Center of Innovation for Complex Chronic Healthcare (CINCCH), Edward Hines, Jr. VA Hospital, 5000 S 5 Ave, Hines, IL, 60141, USA; Peter O’Donnell Jr. School of Public Health, UT Southwestern Medical Center, 5323 Harry Hines Blvd, Dallas, TX 75390, USA; Department of Neurology, Edward Hines, Jr. VA Hospital, 5000 S 5 Ave, Hines, IL, 60141, USA; Department of Physical Medicine and Rehabilitation, Northwestern University, Feinberg School of Medicine, 250 E Superior St, Chicago, IL 60611, USA; Cooperative Studies Program, Edward Hines, Jr. VA Hospital, 5000 S 5 Ave, Hines, IL, 60141, USA; Division of Epidemiology and Biostatistics and Department of Psychiatry, University of Illinois at Chicago, 601 S Morgan St, Chicago, IL 60607, USA; Department of Psychiatry and Behavioral Sciences, Northwestern University, Feinberg School of Medicine, 250 E Superior St, Chicago, IL 60611, USA

**Keywords:** neuromodulation, transcranial magnetic stimulation, intermittent theta burst stimulation, chronic pain, yoga

## Abstract

The complex clinical presentation of patients with mild traumatic brain injury and chronic pain (mTBI+CP) often causes symptoms that are resistant to standard treatments. For the first time, our team combined intermittent theta burst stimulation and yoga (iTBS+LYB) to maximize treatment effect and promote functional recovery in a pilot open-label clinical trial among Veterans with mTBI+CP. A 3-minute iTBS protocol was delivered including 3 pulses of stimulation given at 50Hz, repeated every 200ms at 80% of the resting MT, with an inter-pulse-interval of 20ms. A 2s train of iTBS was repeated every 10s for a total of 190s, for a total of 600 pulses. After all participants received iTBS, a certified yoga instructor guided a 90-minute group LYB session. N=19 participants were enrolled; 71.4% (10/14) completed all 6 iTBS+LYB sessions. Three individuals experienced headaches, a known side effect of iTBS, which resulted in one participant self-withdrawing. iTBS+LYB for mTBI+CP is feasible and safe, with known side effects.

---

More than half of individuals with mild traumatic brain injury (mTBI) have comorbid chronic pain (CP). The complex clinical presentation of such patients often causes symptoms that are resistant to standard treatments. Types of transcranial magnetic stimulation (TMS)^1^, including intermittent theta burst stimulation (iTBS)^2^, applied to the motor cortex (MC) are known to improve pain outcomes. iTBS is a type of patterned, excitatory TMS that enhances MC excitability for at least 60 minutes,^3^ thus also enhancing the impact of interventions provided during this window.^4^

A meta-analysis on the effects of mindfulness-based interventions, including yoga, for mild TBI (mTBI) reveals significant pain and quality of life (QOL) improvements relative to controls.^5^ LoveYourBrain Yoga (LYB), specifically created for individuals with TBI, improves QOL.^6^ For the first time, our team combined, iTBS and yoga (iTBS+LYB) to maximize treatment effect and promote functional recovery in a pilot open-label clinical trial among Veterans with mTBI+CP (ClinicalTrials.gov ID: NCT04517604).

Procedures were completed in accordance with an IRB-approved protocol and an FDA Investigational Device Exemption (G200195). All participants provided informed consent. Prior to intervention administration, participants met with a physiatrist to determine if mat versus chair yoga would be appropriate and review their current pain management strategies. Full methods are published elsewhere;^7^ eligibility criteria are reported in Supplement 1. All participants received iTBS+LYB. mTBI was defined using the Symptom Attribution and Classification Algorithm (SACA), implementing VA/DoD criteria for identifying mTBI. Chronic pain was defined as chronic musculoskeletal pain that persists for >6 months of moderate-to severe-intensity based on patient-reported item scores on the Brief Pain Inventory (BPI).

Stimulation intensity for treatment was determined using each participant’s resting motor threshold (MT), defined as the lowest intensity necessary to produce motor-evoked potentials (MEPs) ≥ 50µV in 5/10 trials.

Provision of all stimulation was completed using a Localite TMS Neural Navigator system using each participant’s T1 MPRAGE. A MagVenture C-B60 coil was used to deliver single-pulse TMS to the dominant MC representation of the abductor pollicis brevis muscle. The treatment stimulation site, the representation of the trunk in the dominant MC, was localized using a T1 MRI via neuronavigation (TMS Navigator, Localite, BELG). The trunk stimulation site was chosen based on its superior pain outcome relative to stimulating the DLPFC among patients with CP receiving TMS.^8^ This site was identified using anatomical landmarks by consensus between the PI and the study neuroradiologist. The neuroradiologist reviewed the T1 and T2 FLAIR for exclusionary abnormalities. An onsite on-call physician was available during each iTBS treatment session to consult with the PI on safety indicators changed from baseline to determine safety to proceed with stimulation. A three-minute iTBS protocol was delivered utilizing the Mag-Pro X100 with MagOption stimulator Cool-B60 or equivalent Cool-B65 Butterfly coil (MagVenture, DK). Parameters included 3 pulses of stimulation given at 50Hz, repeated every 200ms at 80% of the resting MT, with an inter-pulse-interval of 20ms. A 2s train of iTBS was repeated every 10s for a total of 190s, for a total of 600 pulses.^9^

Each participant was stimulated in an order largely counterbalanced across 6 sessions (e.g., each participant had equal opportunity to be stimulated 1st across sessions). After all participants received iTBS, a certified yoga instructor guided the group LYB session. This 90-minute session included 10 minutes of breathing exercises, 45 minutes of gentle yoga, 15 minutes of guided meditation, and 20 minutes of facilitated psychoeducation.^6^ A variety of modifications were offered during the gentle yoga portion of the session to ensure safety and comfort based on medical histories, including standing- and seated-based pose options.

N=19 participants were enrolled; 5 were withdrawn prior to intervention due to not meeting eligibility criteria and 14 received at least 1 intervention session (“treatment initiators”). Of treatment initiators, 71.4% (10/14) completed all 6 iTBS+LYB sessions; 3 discontinued due to scheduling issues and 1 self-withdrew (Supplement 2 CONSORT Diagram). Four cohorts comprised of 3 individuals were completed between August 2021–August 2023. Although the enrollment target was 20 participants, given the complexity of establishing cohorts, enrolling a 20^th^ participant was deferred. Notably, our 71% completion rate was higher than the rate seen in the LYB trial (22%) despite the added participant burden of receiving iTBS.^6^ The differences in our research-based versus the community-based setting in which the LYB study was completed may have contributed to these differences. Future publications will examine setting complexities in more depth.

We monitored 17 safety indices before and after each iTBS session: vital signs (temperature, blood pressure, heart rate, oxygen saturation levels), fatigue, tinnitus, sleep, dizziness, nausea, vomiting, confusion, seizure, syncope, headache, neck pain, skin integrity of the scalp, and substance use. We report severity of change from baseline before and after each iTBS session (Table 1). Three individuals experienced headaches, a known side effect of iTBS, which resulted in one participant self-withdrawing. Per participant’s report, the headaches started ∼3 days after the first iTBS+LYB session and were described as right-sided, all-day migraines with photophobia occurring every other day for a month. This participant had history of occasional unlocalized headaches prior to receiving iTBS and severe chronic pain, primarily in the lower back. TMS has been recommended by an expert consensus panel as a clinical treatment option for post-TBI headaches,^10^ and has been explored as a potential preventive treatment for migraine.^10^ The chronology of this participant’s symptoms and the laterality being opposite of the stimulated side is atypical with how post-TMS-associated headaches have been described.^9^ However, we cannot rule out that post-iTBS (vs post-TMS) headaches may present differently; most of the post-TMS-associated headache literature does not specifically include iTBS protocols. It will be important to continue to follow headache side effects in future larger-scale iTBS+LYB studies.

**Table 1.**
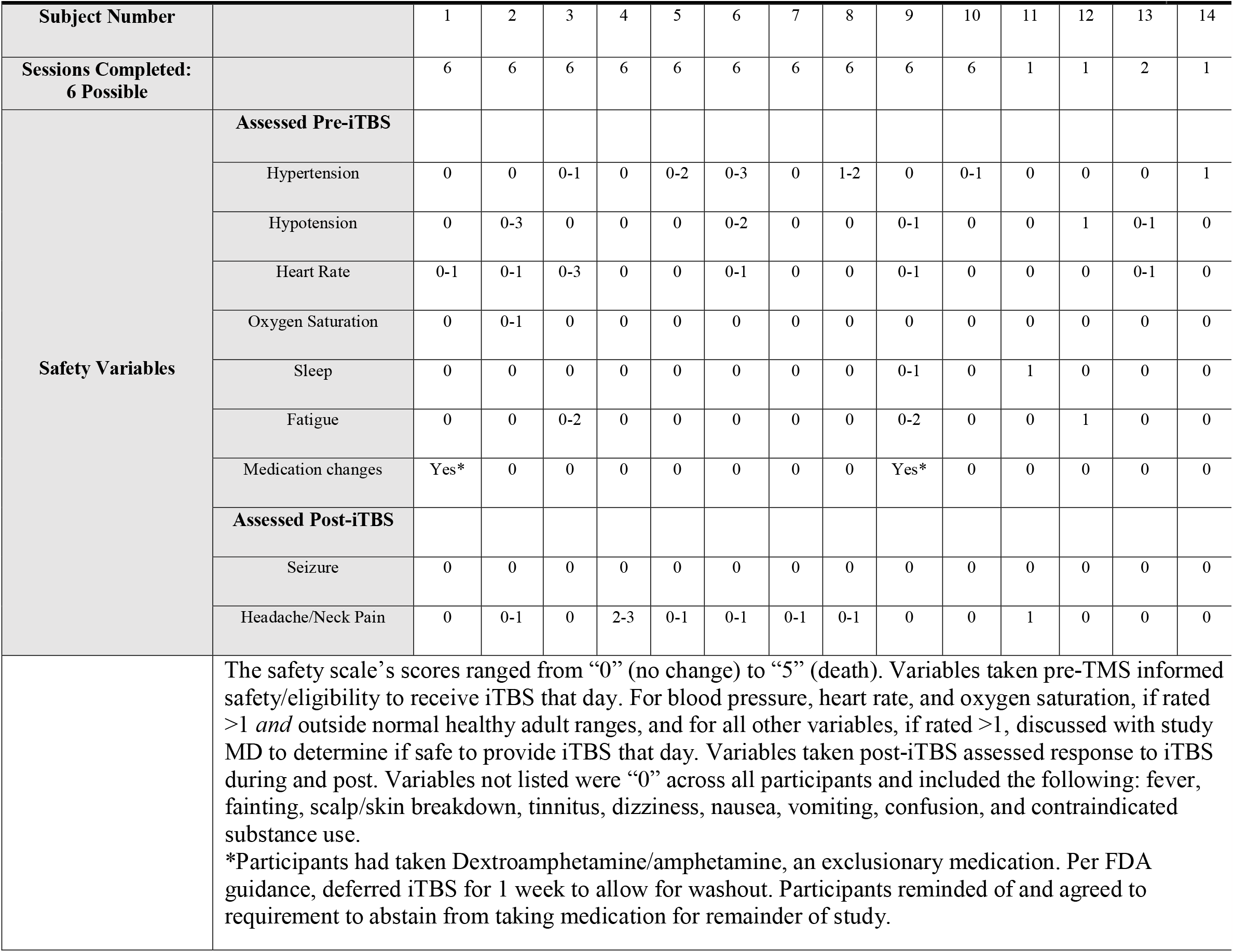
Range of Safety Severity Ratings across iTBS Sessions.

Our findings suggest that administering iTBS+LYB for mTBI+CP is feasible and safe, with known TMS side effects. Limitations of these findings include the small sample size and lack of a control group. Future studies will include a larger sample, as well as the following additional groups: (1) sham-iTBS+yoga, (2) iTBS alone, and (3) treatment as usual.

## Declarations of Interest

KD is employed by the LoveYourBrain Foundation, a nonprofit, for whom she led the design of the curriculum for the LoveYourBrain Yoga program. KD is married to the Executive Director of the LoveYourBrain Foundation. KK is a volunteer for the LYB Foundation, serving as a “Clinical Connector” to raise awareness amongst patients about the opportunities LYB offers. AA, RS, SC, SB, BE, SB, IK, AB, KC, SK, TLP, and DB have no relationships or activities that could appear to have influenced the submitted work. The authors alone are responsible for the content and writing of the paper.

## Disclosures

All data presented in this Letter to the Editor are original contributions and are not published elsewhere.

- Prior presentation of this preliminary safety data: Krese KA, Donnelly KZ, Etingen B, Billups A, Herrold AA. Preliminary results from an open label, pilot, combined neuromodulation and yoga intervention study for mild TBI and chronic pain. American college of Rehabilitation Medicine (ACRM). Chicago, IL. November 11, 2022.
- An a priori manuscript describing the protocol of this project was published in 2022: Krese KA, Donnelly KZ, Etingen B, Bender Pape TL, Chaudhuri S, Aaronson AL, Shah RP, Bhaumik DK, Billups A, Bedo S, Wanicek-Squeo MT, Bobra S, Herrold AA. Feasibility of a Combined Neuromodulation and Yoga Intervention for Mild Traumatic Brain Injury and Chronic Pain: Protocol for an Open-label Pilot Trial. JMIR Res Protoc. 2022 Jun 15;11(6):e37836. doi: 10.2196/37836. PMID: 35704372; PMCID: PMC9244651.

## Data Availability

All deidentified data produced in the present study are available upon reasonable request to the authors.

## Acknowledgements

The authors gratefully acknowledge the LoveYourBrain foundation, Hines VA TBI/polytrauma and pain clinics, the Hines Veterans Engagement Panel, and the Shirley Ryan AbilityLab Pain Management Center.

## Funding support

The study funded by a VA Rehabilitation Research & Development Small Projects in Rehabilitation Research (SPiRE) grant which began on 12/1/2020 (I21RX003611) and the Edward Hines Jr., VA CARES SERWA grant.

**Table 1:** Safety Events

**Supplement 1:** Eligibility criteria

**Supplement 2**: CONSORT diagram

## Notes

### Clinical Trial

NCT04517604

### Clinical Protocols

https://www.researchprotocols.org/2022/6/e37836

### Author Declarations

IRB of Edward Hines, Jr VA Hospital gave ethical approval for this work

