## Supplemental File 2 for "Safety and feasibility of a combined neuromodulation and yoga intervention for individuals with mild traumatic brain injury and chronic pain: a pilot study"

**Supplement** **2**: **CONSORT Flow Diagram**. Number of participants at each study phase are illustrated below.

**Enrollment**

**Intervention**

**Follow-Up**

**Analysis**

**Cohort**

**Assignment**

Consented & completed in person eligibility screening (n= 19)

Excluded (n= 4)

♦  Not meeting inclusion criteria (n= 3)

♦	Participant schedule changes (n=1)

Lost to follow-up (n= 0)

Received intervention (n= 14)

♦ Intervention completers (n= 10)

♦ Intervention non-completers (n=4)

Analysed (n= 14)
♦ All participants who completed at least one iTBS session were included in safety analysis

Discontinued after 1-2 sessions (n= 4)

♦  Participant schedule changes (n= 3)

♦  Self-withdrew due to headaches (n= 1)

Cohort 1 (n=3)

Cohort 2 (n=3)

Cohort 3 (n=3)

Cohort 4 (n=3)

Cohort 5 (n=3)

Excluded (n=1) new contraindication to iTBS; positive drug test
