## Supplemental File 1 for "Safety and feasibility of a combined neuromodulation and yoga intervention for individuals with mild traumatic brain injury and chronic pain: a pilot study"

**Supplement 1. Study Eligibility.** Inclusion and exclusion criteria for participation in the study are listed below.

| **Inclusion Criteria** |
| --- |
| - 22+ years of age - Can read and speak English - Perceive themselves as able to participate in gentle physical movements and cleared by study physician to do so. - mTBI Criteria: Symptom Attribution and Classification (SACA) criteria for mTBI (without requirement of clinical neuropsychological impairment) - Chronic pain: pain (in muscles, bones, ligaments, tendons and/or nerves) that persists for >6 months and is of moderate to severe intensity with a score of >5 on specific items on the Brief Pain Inventory (BPI) - Fully vaccinated against COVID-19 prior to study participation |
| **Exclusion Criteria** |
| - Contraindications to iTBS/TMS (e.g. epilepsy, history of anoxic brain injury or heart disease) - Contraindications to MRI (e.g., claustrophobia, ferromagnetic metal implants) - Pain believed to be associated with cardiac or ischemic conditions - History of moderate to severe TBI - Active seizure disorder, or if they are taking psychostimulants (e.g. amphetamines), anticholinergics or other medications that may increase their risk of having seizures - History of or current psychosis not due to an external cause (e.g., due to illicit drug use) - Are pregnant or nursing - Within 12 weeks of a major surgery/operation - Have questionably valid test profiles |
